# Best practices for multi-ancestry, meta-analytic transcriptome-wide association studies: lessons from the Global Biobank Meta-analysis Initiative

**DOI:** 10.1101/2021.11.24.21266825

**Authors:** Arjun Bhattacharya, Jibril B. Hirbo, Dan Zhou, Wei Zhou, Jie Zheng, Masahiro Kanai, the Global Biobank Meta-analysis Initiative, Bogdan Pasaniuc, Eric R. Gamazon, Nancy J. Cox

**Affiliations:** Department of Pathology and Laboratory Medicine, David Geffen School of Medicine, University of California, Los Angeles, CA, United States; Institute of Quantitative and Computational Biosciences, David Geffen School of Medicine, University of California, Los Angeles, CA, United States; Department of Medicine, Division of Genetic Medicine, Vanderbilt University School of Medicine, Nashville, TN, United States; Vanderbilt Genetics Institute, Vanderbilt University Medical Center, Nashville, TN, United States; Analytic and Translational Genetics Unit, Massachusetts General Hospital, Boston, MA; Program in Medical and Population Genetics, Broad Institute of Harvard and MIT, Cambridge, MA; Stanley Center for Psychiatric Research, Broad Institute of Harvard and MIT, Cambridge, MA; MRC Integrative Epidemiology Unit (IEU), Bristol Medical School, University of Bristol, Oakfield House, Oakfield Grove, Bristol, BS8 2BN, UK; Department of Biomedical Informatics, Harvard Medical School, Boston, MA, USA; Department of Statistical Genetics, Osaka University Graduate School of Medicine, Suita 565-0871, Japan; Department of Human Genetics, David Geffen School of Medicine, University of California, Los Angeles, CA, United States; Department of Computational Medicine, David Geffen School of Medicine, University of California, Los Angeles, CA, United States; MRC Epidemiology Unit, University of Cambridge, Cambridge, UK

**Keywords:** transcriptome-wide association study, meta-analysis, multi-ancestry genetic analysis, Global Biobank Meta-analysis Initiative

## Abstract

**SUMMARY:** The Global Biobank Meta-analysis Initiative (GBMI), through its genetic and demographic diversity, provides a valuable opportunity to study population-wide and ancestry-specific genetic associations. However, with multiple ascertainment strategies and multi-ethnic study populations across biobanks, the GBMI provides a distinct set of challenges in implementing statistical genetics methods. Transcriptome-wide association studies (TWAS) are a popular tool to boost detection power for and provide biological context to genetic associations by integrating single nucleotide polymorphism to trait (SNP-trait) associations from genome-wide association studies (GWAS) with SNP-based predictive models of gene expression. TWAS presents unique challenges beyond GWAS, especially in a multi-biobank and meta-analytic setting like the GBMI. In this work, we present the GBMI TWAS pipeline, outlining practical considerations for ancestry and tissue specificity and meta-analytic strategies, as well as open challenges at every step of the framework. Our work provides a strong foundation for adding tissue-specific gene expression context to biobank-linked genetic association studies, allowing for ancestry-aware discovery to accelerate genomic medicine.

## INTRODUCTION

Large population-based or clinical-case based biobanks are a key component of precision medicine efforts and provide opportunities for genetic and genomic research (Abul-Husn and Kenny, 2019). Biobanks offer context to deploy genome-wide associations (GWAS) at scale. Multi-biobank collaborations facilitate well-powered, multi-ethnic genetic research (Swede et al., 2007). In addition, such collaborations can accelerate the elucidation of the biological mechanisms that underlie diseases by in-silico longitudinal genetic studies and examination of pleiotropy.

A key challenge in GWAS is interpreting significant trait-associated loci and these loci to genes or epigenomic features (Gallagher and Chen-Plotkin, 2018; Wijmenga and Zhernakova, 2018). Viable options to add biological interpretation to our understanding of GWAS loci include colocalization (Giambartolomei et al., 2014, 2018; Gleason et al., 2020; He et al., 2013) or Mendelian randomization methods (Hauberg et al., 2017; Pavlides et al., 2016; Smith and Ebrahim, 2003). Another suite of tools include transcriptome-wide association studies (TWAS), which integrate GWAS with expression quantitative trait loci (eQTL) analyses to prioritize gene-trait associations (GTAs) with applications of mediation analysis (Gamazon et al., 2015; Gusev et al., 2016) or Mendelian randomization (Zhang et al., 2020). TWAS involves three general steps. First, per-gene predictive models of gene expression are trained in the eQTL dataset using genetic variants. Then, genetically-regulated expression (GReX) is imputed in the GWAS cohort with individual-level genotypes. Lastly, statistical associations between GReX and trait are estimated (Barbeira et al., 2018; Gamazon et al., 2015; Gusev et al., 2016). TWAS is also viable with GWAS summary statistics by estimating the test statistic of the TWAS association using a proper LD reference panel (Gusev et al., 2016). Generally, most TWAS methods predict expression using SNPs local to the gene within 1 Megabase of the gene body (Barbeira et al., 2018; Gamazon et al., 2015; Gusev et al., 2016; Hu et al., 2019; Nagpal et al., 2019; Parrish et al., 2022; Zhou et al., 2020). Recently, methods that include strong distal-eQTL signals have shown improved prediction and power to detect GTAs (Bhattacharya et al., 2021a; Luningham et al., 2020). Nonetheless, practical and statistical considerations to accurately prioritize GTAs through TWAS still require methodological improvement.

Along with those from traditional GWAS, TWAS introduces new challenges by incorporating gene expression (Wainberg et al., 2019) (**Figure 1A**). On the genetic level, as in GWAS, disentangling signals from complex LD structure, relatedness, and ancestry requires careful modeling considerations (Mbatchou et al., 2020; Zhou et al., 2018). Selection of LD reference is specifically important in multi-ancestry settings, like the GBMI, as LD structure across ancestry groups differs greatly(Shifman et al., 2003). Mismatched LD may lead to gene expression models with reduced predictive power, reduced power to detect GTAs, and increased false positives (Bhattacharya et al., 2020; Geoffroy et al., 2020; Keys et al., 2020). In addition, phenotype acquisition and aggregation are challenging, especially across multiple biobanks with different healthcare, electronic health record, and case-control definitions. However, compared to GWAS, a challenge specific to TWAS is the integration of gene expression with GWAS signal. Not only is it an active topic of methodological research to choose an optimal set of genes and tissues that best explains the phenotype association at a given genetic locus, the role of context-specific expression is still being evaluated in trait associations. Dynamic differences in bulk tissue expression from cell-type- or cell-state-specificity can give additional granularity to gene-trait associations. The impact of these challenges in a meta-analytic framework has not been previously explored.

**Figure 1:**
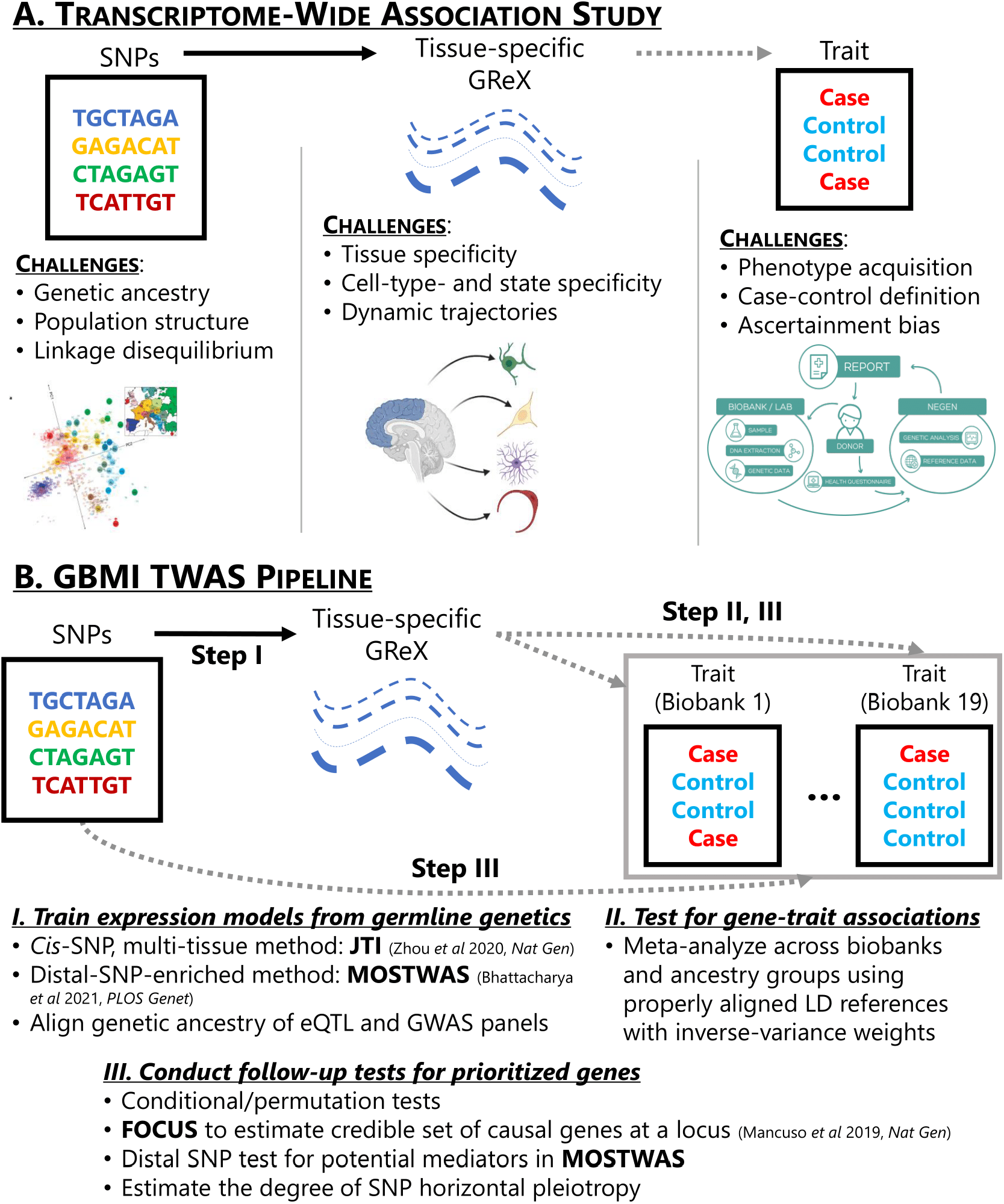
Overview of GBMI transcriptome-wide association study (TWAS) pipeline with challenges at every data level. **(A)** Each level of data in TWAS introduces a unique set of challenges: (1) genetics data include confounding from genetic ancestry, population structure and relatedness, and complex linkage disequilibrium patterns, (2) gene expression data introduces context-specific factors, such as tissue-, cell-type-, or cell-state-specific expression, and (3) phenotypic data, especially in the meta-analyses of multiple biobanks, involve challenges in acquiring and aggregating phenotypes, properly defining controls for phenotypes, and ascertainment and selection bias from non-random sampling. **(B)** An overview of the GBMI TWAS pipeline: (1) JTI and MOSTWAS for model training, (2) inverse-variance weighted meta-analysis using per-biobank, per-ancestry group TWAS summary statistics, and (3) various follow-up tests, including conditional or permutation tests, distal-SNPs added last test, probabilistic fine-mapping using FOCUS, and tests for SNP horizontal pleiotropy. Dotted lines represent associations that are tested in the TWAS pipeline, while the solid lines represent a link built through predictive modeling.

Here, we outline a framework for analyzing trans-ancestry, meta-analytic GWAS across multiple biobanks with TWAS. We review and explore practical considerations for all three steps (**Figure 1B**): ancestry specificity of expression models and LD reference panels, meta-analytic techniques for detection of GTAs, and follow-up tests and analyses for biological context. Our framework can be applied to various phenotypes to study population-wide and ancestry-specific genetic associations mediated by tissue-specific expression.

## RESULTS

### Expression models are not portable across ancestry groups

The diversity represented in the GBMI enables uniquely well-powered studies to detect genetic associations in non-European populations. However, optimal TWAS requires ancestry-matched training datasets of genetic and tissue-specific gene expression data, which are still lacking in non-European samples. As Cao *et al* points out (Cao et al., 2021), statistical power to detect GTAs in TWAS is dependent on expression heritability and the ability of the predictive expression model to recapitulate that heritable expression in the external GWAS panel. Accordingly, training expression models that perform well in all ancestry populations is necessary to ensure that discoveries made through TWAS are not restricted to European populations. For the first GBMI TWAS, we restrict analysis to populations of European ancestry, due to small sample sizes of non-European ancestry samples (Aguet et al., 2020). However, as sample sizes for eQTL datasets in non-European populations increase, the TWAS pipeline will include expression models for these understudied and underserved populations (**STAR Methods**). Here, we illustrate some challenges in building these expression models across ancestry groups.

We considered the 5 tissues in GTEx with at least 70 samples from both European (EUR) and African (AFR) ancestry: subcutaneous adipose (abbreviated ADIP, *N*=*71* samples of AFR ancestry and 492 samples of EUR ancestry), tibial artery (ARTERY, *N*=*76* and 489), skeletal muscle (MUSC, *N*=*86* and 602), sun exposed lower leg skin (SKIN, *N*=*73* and 518), and whole blood (BLOOD, *N*=*80* and 574). For genes with significantly heritable expression in both EUR and AFR GTEx samples (restricted maximum likelihood-based estimate of heritability >*0* with nominal *P <0*.*01*), we trained EUR- and AFR-specific models using elastic net regularized regression (Friedman et al., 2010) and imputed expression into the aligned (i.e., training and imputation samples have similar ancestries) and misaligned (i.e., training and imputation sample have different ancestries). For context, we also built ancestry-unaware models, where EUR and AFR samples were pooled together. We calculated predictive performance with adjusted R^2^ to account for sample size, using leave-one-out CV when measuring predicting performance in an aligned imputation sample (**STAR Methods**).

Across these tissues, models trained in EUR samples performed, on average, 4 times worse (differences of 0.03-0.04 in median adjusted R^2^) in AFR samples compared to models trained in AFR samples (**Figure 2A, Tables S1-S3**), with more than 80% of gene models with stronger performance if trained in AFR samples. Similar trends hold for ancestry-specific models imputed into down-sampled EUR imputation samples (**Figure S1-S2, Table S1-S3**), consistent with previous simulation and real-world studies (Bhattacharya et al., 2020; Keys et al., 2020); here, we considered a randomly selected EUR imputation sample with equal sample size to that of the AFR sample in the same tissue. In fact, we observed that ancestry-specific models imputed into a sample with aligned ancestry showed larger predictive R^2^ than ancestry-unaware (individuals of EUR and AFR ancestry in the training sample) models imputed into the same sample (**Figure 2B, Table S4**), despite generally increased sample sizes. This observation also holds if we further increase the sample size of the training sample by including individuals of other ancestries (Asian, American Indian, and Unknown ancestries) into the training sample (**Figure S3**). This observation emphasizes the need for ancestry matching in gene prediction from genetic data and greater recruitment of non-European ancestry patients in eQTL studies.

**Figure 2:**
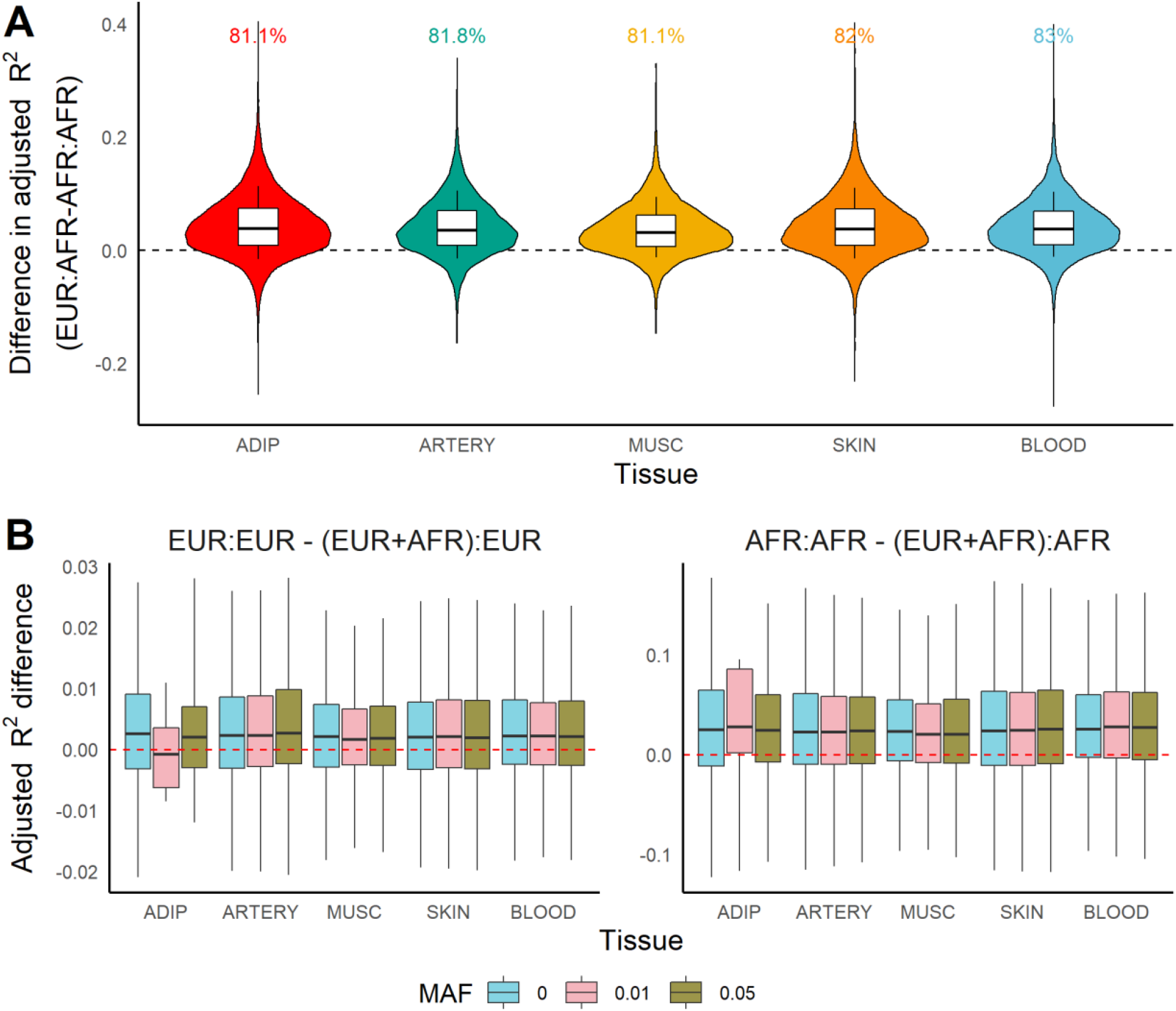
Comparison of predictive performance of genetic models of expression across ancestry. **(A)** Distribution of difference in adjusted R^2^ (Y-axis) when predicting expression in the AFR imputation sample between models trained in EUR and in AFR training samples across tissue (X-axis). **(B)** Distribution of difference in adjusted R^2^ between ancestry-specific and ancestry-unaware models imputing into EUR (left) and AFR (right) samples.

In these analyses, one reason ancestry-unaware models may perform poorly in AFR samples is due to differences in minor allele frequency (MAF) of highly predictive SNPs between EUR and AFR ancestry populations. It is important to note that this discrepancy is not generally specific to any one ancestry; rather, ancestry imbalance in the training or reference datasets may lead to poor portability of genetic models due to differences in allele frequency. To account for common SNPs in both AFR or EUR ancestry populations, we additionally trained ancestry-unaware and ancestry-specific models using SNPs with minor allele frequency (MAF) exceeding various thresholds in both AFR and EUR samples. Excluding SNPs with MAF < 0.01 improved predictive performance of ancestry-unaware models across all tissues (**Figure S4, Table S5**). However, the gap in predictive performance between ancestry-specific and ancestry-unaware models did not decrease when the MAF cutoff was increased (**Figure 2B, Table S4**). This observation may reflect that dropping ancestry-specific rarer SNPs ignores variants with large ancestry-specific effects on gene expression. Additionally, excluding rare ancestry-specific SNPs does not address the differences in LD across the EUR and AFR samples which leads to different regularization paths and, hence, SNP-gene weights. Addressing the trans-ancestry portability of expression models remains an open study direction; methodology that borrows information from functional annotations or across different cell-type- or cell-state-specific contexts may bridge this gap in predictive performance, similar to recent developments in polygenic risk score prediction for complex traits (Amariuta et al., 2020; Márquez-Luna et al., 2020).

### Meta-analytic strategies must be ancestry-aware

Another critical consideration for the GBMI involves meta-analysis while using GWAS summary statistics. TWAS estimates the association between GReX and the phenotype by weighting the standardized SNP-trait effect sizes from GWAS summary statistics by SNP-gene weights from the expression models. To account for the correlation between SNPs, an external LD reference panel, like the 1000Genomes Project (Auton et al., 2015), is used to estimate the standard error of the TWAS association. Accordingly, the accuracy of this reference panel to the LD structure in the GWAS cohort dictates how aligned the summary-statistics based TWAS association is to the TWAS association from direct imputation into individual-level genotypes in the GWAS cohort. Ideally, in-sample LD will give the best estimate of the TWAS standard error, but several biobanks do not provide this information under their specific genetic data sharing and privacy policies. Even departures in LD across subgroups of European ancestry populations may influence the standard error estimate. In addition, as the estimates of SNP-gene weights are influenced by the LD in the eQTL panel, differences in LD between the eQTL and GWAS panel will also affect the TWAS effect size.

As LD structure greatly differs across ancestry groups (Shifman et al., 2003), pooling ancestry groups in TWAS may lead to reduced power. We conducted TWAS for asthma risk using ancestry-unaware and EUR- and AFR-specific models of whole blood expression (4,782 genes with heritable expression at nominal P < 0.01 and models with cross-validation R^2^ > 0.01 with nominal P < 0.05, trained via elastic net regression). Ancestry-specific TWAS Z-scores across EUR and AFR ancestry groups were not strongly correlated (*r*=*0*.*11*), potentially due to differences in sample size and eQTL and GWAS architecture (**Figure 3A, Figure S5-S6**) (Shang et al., 2020; Wyss et al., 2018). In fact, we detected only two genes across both EUR and AFR with P < *2*.*5* × *10*^−*6*^. One of these genes, *DFFA*, has been implicated with asthma risk through GWAS and colocalization in EUR (Vicente et al., 2017). However, the TWAS associations across EUR and AFR were in opposite directions using blood tissue. In the other 4 tissues explored, *DFFA* TWAS associations did not reach transcriptome-wide significance but effect directions were generally concordant (**Figure S7**). In blood, lead local-eQTLs (within 1 Megabase) of *DFFA* show are in opposite directions, though only nominally significant at P < 0.05 (**Figure S8**). Although they are within 60 kilobases, the lead eQTLs for DFFA across AFR (rs263526) and EUR (rs903916) are not in LD (R^2^ = 3 × 10^−4^ in AFR, 0.0072 in EUR). The GWAS effect sizes of SNPs local to *DFFA* do not show large deviations in effect direction and are only nominally significant, as well (**Figure S8**). These differences in TWAS associations across ancestry motivate careful consideration of meta-analytic strategy to avoid biasing cross-ancestry associations towards cohorts with larger sample sizes, which still tend to be predominantly of EUR ancestry.

**Figure 3:**
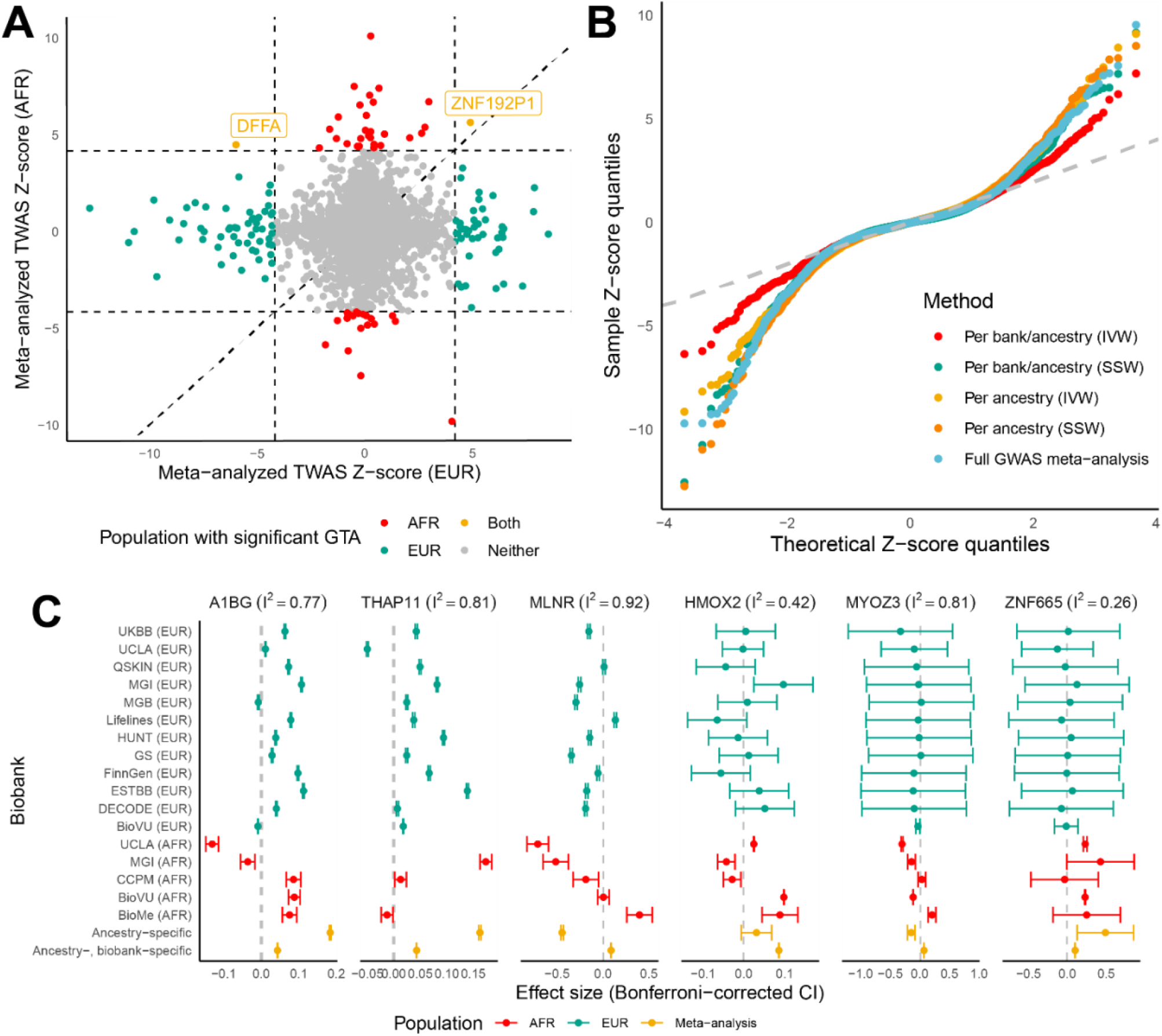
Comparison of meta-analytic strategies for multi-biobank, trans-ancestry TWAS. **(A)** Scatterplot of per-ancestry meta-analyzed TWAS scores across EUR (X-axis) and AFR ancestry (Y-axis). The dotted horizontal and vertical lines indicate P < *2*.*5* × *10*^−*6*^ with a diagonal line for reference. Points are colored based on which ancestry population the TWAS association meets P < *2*.*5* × *10*^−*6*^. **(B)** QQ-plot of TWAS Z-scores, colored by meta-analytic strategies. Per ancestry refers to TWAS meta-analysis across meta-analyzed ancestry-specific GWAS summary statistics. Per bank/per ancestry refers to TWAS meta-analysis using all biobank- and ancestry-specific GWAS summary statistics. **(C)** Effect sizes and Bonferroni-corrected confidence intervals (CIs) for TWAS associations across 17 individual biobanks (stratified by ancestry group with EUR in green and AFR in red) and 2 IVW meta-analysis strategies (in yellow) for 5 representative genes. The Higgins-Thompson I^2^ statistic for heterogeneity is provided, with the dotted line showing the null.

We investigated 5 different meta-analytic strategies empirically: meta-analyzing across ancestry-specific, per-biobank GWAS summary statistics using (1) inverse-variance weighting (IVW) and (2) sample-size weighting (SSW), meta-analyzing across ancestry-specific meta-analyzed GWAS summary statistics using (3) IVW and (4) SSW, and (5) TWAS using ancestry-unaware models and meta-analyzed GWAS summary statistics across EUR and AFR ancestry groups (**STAR Methods**). QQ-plots in **Figure 3B** show earlier departure of Z-scores from the QQ-line for SSW meta-analyzed Z-scores and the ancestry-unaware strategy, suggesting inflation. This observation is supported with estimates of test statistic bias and inflation using an empirical Bayes method, bacon (van Iterson et al., 2017), which show the largest estimated bias and inflation for these SSW and ancestry-unaware methods. IVW strategies show similar levels of inflation, with IVW meta-analysis across ancestry-specific meta-analyzed GWAS summary statistics showing minimal bias (**Figure S9**). These results align with intuition – that the more naïve SSW meta-analysis and ancestry-unaware methods bias towards EUR cohorts, which have the larger sample sizes, whereas Z-scores from the IVW methods showed positive correlations with Z-scores from AFR cohorts (**Figure S6**).

However, it is unclear whether ancestry-specific IVW meta-analysis to the per-biobank level is necessary. As shown in **Figure S10**, Z-scores from these two IVW methods are moderately positively correlated (*r* = *0*.*51* across 4,152 Z-scores), with this correlation increasing when we consider genes with nominally significant Z-scores for both strategies (*r*=*0*.*70* across 564 tests). We observed that top associations across these IVW meta-analyses often had high degrees of heterogeneity in effect size across biobanks, as measured by the Higgins-Thompson I^2^ statistic (**Figure 3C, Figure S11**) (Higgins and Thompson, 2002). One gene, *A1BG*, that showed directionally concordant transcriptome-wide significant associations across both IVW strategies had a large degree of heterogeneity in the underlying participating cohorts (I^2^= 0.77). In fact, the cross-biobank heterogeneity is often larger than the cross-ancestry heterogeneity for TWAS associations of *A1BG*. Interestingly, *ZNF665*, another gene with directionally concordant associations across both IVW strategies showed a low degree of heterogeneity in the per-biobank effect sizes (I^2^ = 0.26). However, genes with discordant associations across IVW strategies showed varied patterns. Two illustrative examples are *MLNR* and *MYOZ3*, both with large degrees of test statistic heterogeneity (I^2^ = 0.91 and 0.82, respectively). Across the two IVW strategies, effect sizes are in opposite directions, possibly due to large standard error differences across the ancestry-specific per-biobank associations. A thorough investigation of the power and false discovery rates of these meta-analysis strategies through simulations is necessary. More sophisticated methods (Hedges and Vevea, 1998; Lee et al., 2017; Shi and Lee, 2016) that can properly incorporate the per-biobank uncertainty into meta-analyzed TWAS associations must be explored to increase power and properly leverage the large sample sizes of the GBMI.

In addition to considerations for TWAS in trans-ethnic populations, analyzing genetic data from individuals of admixed ancestry is also an open area of study. For example, in this analysis, we have used the 1000Genomes AFR LD reference panel as an estimate of the LD for the AFR-ancestry samples from each biobank. However, most of these AFR-ancestry populations are of admixed ancestry (e.g, African Americans or African British). A single LD reference panel of AFR-ancestry may not reflect the genetic diversity in these admixed populations of AFR and EUR ancestries from around the world (Baharian et al., 2016). As Zhong et al highlights, in multiethnic and admixed populations, using local ancestry estimates aids in better characterization of heritability of complex traits and more accurate mapping of genetic associations, especially eQTLs (Zhong et al., 2019). Accordingly, incorporating local ancestry estimates into both expression model step and the association testing step of TWAS may lead to increased power and should be explored.

### Follow-up tests provide biological and clinical context to TWAS GTAs

TWAS GTAs identified using GWAS summary statistics are subject to several factors that may lead to false positives. We implement several follow-up tests to provide context to TWAS-identified GTAs. First, a TWAS GTA could attain transcriptome-wide significance due to only the strong SNP-trait associations from the underlying GWAS. To quantify the significance of the GTA conditional on the SNP-trait effects at the locus, we perform a permutation test by permuting the SNP-gene weights from the expression model to generate a null distribution (**STAR Methods**). Comparing the original TWAS Z-score to this null distribution assesses how much signal is added by the expression given the specific GWAS architecture of the locus. As Gusev *et al* point out, this permutation test is highly conservative and intended to prioritize only associations already significant in the standard TWAS GTA detection (Gusev et al., 2016).

Next, gene expression models for genes in adjacent genomic windows may be built from overlapping SNPs or SNPs in strong LD. When TWAS detects GTAs in overlapping genomic regions, we apply Bayesian probabilistic fine-mapping using FOCUS (Mancuso et al., 2019)to estimate a 90% credible set of genes to explain the observed association signal in a given tissue (**STAR Methods**). However, the current iteration of FOCUS has limitations. Priors for the correlation matrix between GReX of overlapping genes are dependent on SNP LD reference panels. Thus, fine-mapping in trans-ancestry settings is difficult, though recent machinery has been added to FOCUS to account for differences in genetic architecture across the study sample (Gopalan and Lu et al, in preparation). Another challenge for gene-level fine-mapping in multi-tissue TWAS is distinguishing between overlapping signals across tissues. Primarily due to cross-cell-type variation in expression levels and eQTL architecture, TWAS may prioritize genes in multiple tissues that are overrepresented by the same underlying causal cell-types (Wainberg et al., 2019). This multi-tissue gene prioritization extends to fine-mapping overlapping TWAS signals across tissue, as priors for FOCUS are not tissue-dependent. An interesting future direction involves careful tissue-specific prior elicitation in TWAS fine-mapping – extracting posterior signal that is biologically consistent and meaningful without allowing the prior to dominate.

The GBMI TWAS model incorporates gene expression models using MOSTWAS, a TWAS extension that prioritizes distal-eQTLs by testing their mediation effect through local molecular features (**STAR Methods**). For genes with models trained with MOSTWAS and associated with the trait at transcriptome-wide significance, we test the additional association signal from the distal-SNPs using an added-last test, analogous to a group-added-last test in linear regression (Bhattacharya et al., 2021a). This test also prioritizes sets of genomic or epigenomic features that mediate the predicted distal-eQTLs for subsequent study of upstream, tissue-specific regulation of GTAs. In one application of MOSTWAS, one prioritized functional hypothesis was experimentally validated *in vitro* (Bhattacharya et al., 2021b). As distal-eQTLs are more likely to be tissue- or cell-type-specific (Yang et al., 2017), the association signal from these distal-eQTLs could also be leveraged in cross-tissue fine-mapping strategies.

Lastly, TWAS suffers from severely reduced power and inflated false positives in the presence of SNP pleiotropy, where the genetic variants in the gene expression model affect the trait, independent of gene expression (Veturi and Ritchie, 2018). We encourage estimating the degree of and accounting for SNP pleiotropy using LDA-MR-Egger (Barfield et al., 2018) or PMR-Egger (Yuan et al., 2020), especially in settings with individual-level GWAS genotypes. Applications for these methods using GWAS summary statistics reveals some inflation of standard errors (Zhu et al., 2021), suggesting the need for further evaluation and development of summary statistics-based methods to account for SNP pleiotropy in TWAS GTA detection.

### Biobanks enable GReX-PheWAS for biological context

Biobanks aggregated in the GBMI provide a rich catalog of phenotypes for analysis, with phenotype codes (phecodes) aggregated from ICD codes classified into clinically relevant categories (Wei et al., 2017). This phenotype catalog enables Phenome-Wide Association Studies (PheWAS) as a complement to GWAS by both replicating GWAS associations and providing a larger set of traits associations with GWAS variants. To follow-up on novel TWAS-prioritized genes, we may expand the PheWAS framework to the tissue-specific GReX level in a similarly complementary analysis: GReX-level Phenome-Wide Association Study (GReX-PheWAS), as previously deployed in biobank settings and similar to the PredixVU database (Pathak et al., 2020; Unlu et al., 2019, 2020). Not only do these analyses replicate and detect new TWAS associations, but they can also point to groups of phenotypes that show enrichments for trait-associations for the gene of interest.

We briefly illustrate an example of GReX-PheWAS using 3 genes (**Figure 4, Figure S12-S13, Table S6**): *TAF7*, a novel gene in our TWAS, and *ILRAP18* and *TMEM258*, two genes previously implicated through GWAS (Johansson et al., 2019; Portelli et al., 2020; Reijmerink et al., 2008, 2010; Zhu et al., 2020). These genes were prioritized from European-specific TWAS for asthma risk from Zhou *et al* (Zhou et al., 2021) using lung tissue expression (101,311 cases and 1,118,682 controls): *TAF7* (MOSTWAS model), *IL18RAP* (JTI model), and *TMEM258* (JTI model). European-specific TWAS meta-analysis for asthma detected a negative association with *TAF7* cis-GReX, a gene that did not intersect a GWAS-significant locus. In TWAS follow-up tests, *TAF7* passed permutation testing and was estimated in the 90% credible set at the genomic locus via FOCUS with posterior inclusion probability 1. *TAF7* encodes a component of the TFIID protein complex, which binds to the TATA box in class II promoters and recruits RNA polymerase II and other factors (Bhattacharya et al., 2014). As the clinically-relevant associations for *TAF7* lung GReX are not characterized, we employed GReX-PheWAS in UKBB European-ancestry GWAS summary statistics across 731 traits and diseases with sample sizes greater than 100,000, grouped into 9 categories (**Figure 4, STAR Methods**). We see enrichments for phenotypes of the hematopoietic and musculoskeletal groups (**Figure 4A**) with hypothyroidism and chronic laryngitis as the top phenotype associations (**Figure 4B, Table S6**). These phenotypes include multiple inflammations of organs (e.g., laryngitis, osteitis, meningitis, inflammations of the digestive and respiratory system, etc). We also detected several associations with related respiratory diseases and traits. Similarly, for the two previously implicated genes, we find enrichments for respiratory and hematopoietic GTAs for *ILRAP18* and across multiple categories for *TMEM258*, consistent with the categorized functions and associations of these genes (**Figure S12-S13, Table S6**). The utility of GBMI’s robust roster of phenotypes enables GReX-PheWAS to add biological and clinical context to novel TWAS associations.

**Figure 4:**
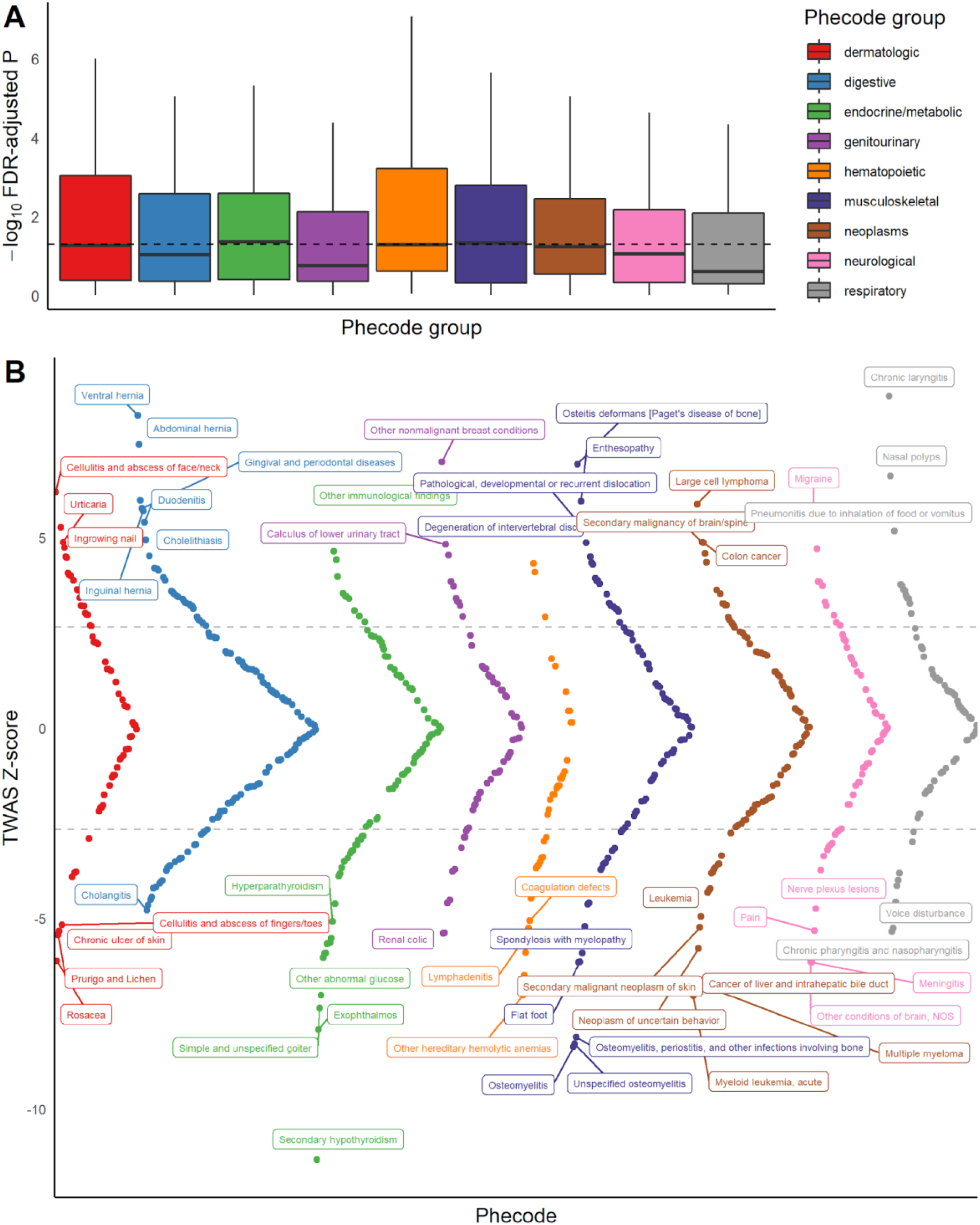
GReX-PheWAS for categorizing phenome-wide associations for TAF7 genetically-regulated expression in UKBB. **(A)** Boxplots of -log_10_ Benjamini-Hochberg FDR-adjusted P-values of GTAs across 9 phenotype groups. The dotted grey line showed FDR-adjusted P = 0.05. **(B)** Miami plot of TWAS Z-scores (Y-axis) across phenotypes, colored by phecode group. The dotted grey line shows the significance threshold for Benjamini-Hochberg FDR correction and phenotypes are labelled if the association passes Bonferroni correction.

GReX-PheWAS, despite its utility, shares the challenges of PheWAS. Phenotypes within and across groups may be correlated, leading to a series of dependent tests. Even divergent phenotypes may be correlated, either clinically or biologically. Thus, simple adjustments of multiple testing burden may not be appropriate, and methods that account for correlation between phenotypes, like permutation tests, may be more applicable (Hebbring, 2014; Korthauer et al., 2019; Stevens et al., 2017; Wei et al., 2017). In addition, covariate adjustments in expression models built for disease-specific analyses may not be generalizable for multiple phenotypes. Most population-based clinical biobanks lack comprehensive clinical and lifestyle information of the individuals. Phenotyping information is typically incomplete, mostly due to gaps in electronic health records. Phenotype groupings may also be deceptive: as most biobanks follow ICD coding that groups traits and diseases by body systems, GReX-PheWAS enrichments for a given group may not reflect shared genetic pathways across body systems. In addition, case-control selection may not be optimal due to differences in exclusion criteria (McGuirl et al., 2020). Lastly, phenotype acquisition and aggregation across multiple biobanks is challenging, with different healthcare, electronic health record, and case-control definition assignments. Despite these limitations in phenotype acquisition, recent methods focusing on identifying shared genetic architecture among multiple phenotypes (McGuirl et al., 2020) in a phenome-wide approach highlight the advantages of GReX-PheWAS.

## DISCUSSION

Here, we provide a framework for TWAS in a multi-biobank setting across many ancestry groups. Our work outlines several methodological gaps that should be addressed in the future: (1) training expression models that are portable across ancestry groups, (2) limiting false discovery in TWAS by properly modeling differences in LD across ancestry groups, (3) incorporating uncertainty within and heterogeneity across biobanks to boost TWAS meta-analytic power, and (4) contextualizing TWAS GTAs through follow-up testing, probabilistic fine-mapping across ancestry groups and expression contexts, and GReX-PheWAS. Along with the discussed issues with current TWAS methodology, tissue-specific expression may not provide sufficient granularity needed to discover trait-relevant biological mechanisms. Recent methods that study the mediation of the SNP-trait relationship by cell-type heterogeneity show that cell-types are influenced by genetics and predict complex traits (Liu et al., 2021). In turn, single-cell eQTL datasets can be integrated with GWAS to identify cell-type- or cell-state-specific expression pathways that are health- or disease-related. Incorporating single cell expression data into a predictive model will require more sophisticated statistical methodology that relies on modeling cell identity as a spectrum, rather than a categorical definition (Burkhardt et al., 2021; Verma and Engelhardt, 2020). Furthermore, a multi-omic approach that incorporates functional data with TWAS may better model the flow of biological information in a biologically interpretable fashion (Baca et al., 2021; Bhattacharya et al., 2021a), with Zhao et al, in preparation.

Despite the limitations of this suite of methods, TWAS continues to be a useful tool for interpreting GWAS associations and independently discovering genetic associations mediated by gene expression. There is a severe need for an increase in reference eQTL data from individuals of non-European ancestry at parity with those of European-ancestry individuals. Moreover, more sophisticated integrative computational and experimental tools to complement improved TWAS and GWAS to understand the biology underlying health and disease need to be developed.

## Data Availability

The all-biobank and ancestry-specific GWAS summary statistics are publicly available for downloading at https://www.globalbiobankmeta.org/resources and browsed at the PheWeb Browser http://results.globalbiobankmeta.org/. 1000 Genome Phase 3 data can be accessed at ftp://ftp.1000genomes.ebi.ac.uk/vol1/ftp/data_collections/1000_genomes_project/data. MOSTWAS can be accessed from https://github.com/bhattacharya-a-bt/MOSTWAS, and JTI can be accessed from https://github.com/gamazonlab/MR-JTI. Sample scripts for this manuscript are available at https://github.com/bhattacharya-a-bt/gbmi_twas.

ftp://ftp.1000genomes.ebi.ac.uk/vol1/ftp/data_collections/1000_genomes_project/data

## ACKNOWLEDGEMENTS

We thank Mark Daly for helpful comments and advice during the internal revision process. We thank Nicholas Mancuso, Michael Love, and Achal Patel for their thoughtful discussion during the research process. We would like to thank the organizing committee of the International Common Disease Alliance for intellectual contributions on the set up of the GBMI as a nascent activity to the larger effort. We would like to thank Daniel King from the Hail team and Sam Bryant from the Stanley Center Data Management team at the Broad Institute for helping with the Google bucket set up and data sharing, and Bethany Klunder from the University of Michigan Medical school for helping with the paper submission.

## AUTHOR CONTRIBUTIONS

Conceptualization: AB, JH; Methodology: AB, JH, DZ, ERG, BP, NJC; Software: AB, JH, DZ, ERG, BP, NJC; Validation: AB, JH, DZ; Formal analysis: AB, JH; Investigation: all authors; Resources: all authors; Data curation: WZ, MK; Writing - original draft: AB, JH; Writing - Review & Editing: all authors; Visualization: AB, JH; Supervision: EG, BP, NJC; Project administration: AB, JH, ERG, BP, NJC; Funding acquisition: all authors

## STAR METHODS

We first outline the steps of the TWAS pipeline employed for phenotype available for analysis in the GBMI. Then, we provide details for the analyses presented in **Results**.

### The GBMI TWAS Pipeline

#### Training expression models from genetics

Tissue-specific expression models trained with reference data from the Genotype-Tissue Expression Project (GTEx) v8 (Aguet et al., 2020) are built using two methods: (1) Joint-Tissue Imputation (JTI), which leverages shared genetic *cis*-regulation across tissues (Zhou et al., 2020), and (2) MOSTWAS, which prioritizes tissue-specific distal-SNPs through rigorous mediation analysis to account for additional expression heritability (Bhattacharya et al., 2021a). Genes with significantly positive expression heritability (nominal *P <0*.*05*) and five-fold cross-validation (CV) adjusted *R*^*2*^ ≥ *0*.*01* with *P <0*.*05* are considered for TWAS. Ancestry-specific models are trained, excluding SNPs with MAF < 0.01 and deviated from Hardy-Weinberg at P < *10*^−*5*^ across all 838 GTEx samples. The first iteration of the GBMI TWAS pipeline focuses on EUR-ancestry models due to larger sample sizes. However, as sample sizes for other ancestry groups increase, this pipeline can be adapted for these currently underrepresented ancestries. In addition, models from other data sources using other methods can be incorporated in subsequent steps.

#### Hypothesis tests for TWAS

To test for an association between tissue-specific GReX of a gene and a trait of interest, GWAS summary statistics are integrated with these expression models. For the EUR-specific TWAS, we use EUR-specific meta-analyzed GWAS summary statistics across all biobanks. JTI and MOSTWAS use two different approaches to test for a GTA. For MR-JTI, the posterior predictive distribution of GReX is estimated, and multiple-instrumental-variable causal inference is used to estimate the GTA, controlling for overall heterogeneity (Zhou et al., 2020). For MOSTWAS, a weighted burden test is constructed, as in FUSION (Bhattacharya et al., 2021a; Gusev et al., 2016; Pasaniuc et al., 2014). Both of these methods require a LD reference panel; the GTEx LD matrix is used as a reference. Taken together, these methods provide effect sizes, standard errors, Z-scores (effect sizes standardized by standard error), and P-values for GTAs. A GTA is transcriptome-wide significant using a Bonferroni correction across all tests run. The number of tests run is equal to the sum of the number of significant gene models across all tissues.

Follow-up tests and analyses are then run to provide context to the TWAS GTAs. A permutation test is run by shuffling the SNP-gene weights 1,000 times and determining the TWAS Z-score at each permutation, generating a null distribution. The original TWAS Z-score is compared to this null distribution to generate a permutation P-value; Benjamini-Hochberg FDR correction is used to account for multiple testing burden here. This test examines whether the SNP-gene relationship provides more information than just the SNP-trait association. Next, for MOSTWAS, the distal-SNPs added-last test is run to measure the association from distal-SNPs in the expression models, conditional on the association from local-SNPs (Bhattacharya et al., 2021a). This test prioritizes sets of mediating molecular features for the SNP-gene relationship with significant effects on the trait. Lastly, for genes whose models are built using SNPs from overlapping genomic regions, probabilistic fine-mapping via FOCUS (default parameters and priors) is employed to determine a 90% credible set of genes that explain the gene-level association signal at the locus (Mancuso et al., 2019). FOCUS also outputs posterior inclusion probabilities for each gene in the 90% credible set.

### Analysis of ancestry-specific and -unaware models

To show the utility of ancestry-specific models, we train EUR- and AFR-specific models using elastic net regression for 5 tissues with more than 70 samples from AFR ancestry patients: subcutaneous adipose (*N*=*492* EUR, *N*=*71* AFR), tibial artery (*N*=*489* EUR, *N*=*76* AFR), skeletal muscle (*N*=*602* EUR, *N*=*86* AFR), sun exposed lower leg skin (*N*=*518* EUR, *N*=*73* AFR), and whole blood (*N*=*574* EUR, *N*=*80* AFR). To balance sample sizes in the imputation sample, we down-sampled the EUR ancestry imputation sample to match the AFR imputation sample. We consider only genes with positive expression heritability in both EUR and AFR training samples (Yang et al., 2011). We also build ancestry-unaware models, where genotypes for EUR and AFR samples are pooled together in the training sample. We calculate predictive performance in aligned and misaligned imputation samples based on ancestry; the aligned imputation sample is one with ancestry that predominantly matches the ancestry of the training sample. Predictive performance is measured with adjusted R^2^ to account for sample size, using an appropriate linear model between predicted and observed expression. For imputation samples that are used in training (aligned imputation panel), we use leave-one-out CV when measuring predictive performance. Lastly, when imputing into AFR and EUR samples using the ancestry-unaware models, we use leave-one-out CV, as well, but only cross-validating over the AFR or EUR samples, respectively.

### Comparison of meta-analytic strategies

We compared 5 different meta-analytic strategies empirically: meta-analyzing across ancestry-specific, per-biobank GWAS summary statistics using (1) inverse-variance weighting (IVW) and (2) sample-size weighting (SSW), meta-analyzing across ancestry-specific meta-analyzed GWAS summary statistics using (3) IVW and (4) SSW, and (5) TWAS using ancestry-unaware models into meta-analyzed GWAS summary statistics across EUR and AFR ancestry groups. First, we consider three different sets of GWAS summary statistics: biobank- and ancestry-specific summary statistics, ancestry-specific summary statistics meta-analyzed across all biobanks, and summary statistics meta-analyzed across biobanks and ancestry groups. In two former settings, for biobank *i* and a given gene, we generate *β*_*TWAS,i*_, the TWAS effect size, and *SE*_*TWAS,i*_, the corresponding standard error. Given *B* different biobanks, the IVW TWAS Z-score, *Z*_*TWAS,IVW*_, is calculated as:

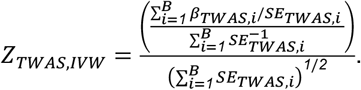

With *Z*_*TWAS,i*_=*β*_*TWAS,i*_/*SE*_*TWAS,i*_ and *N*_*i*_ as the sample size of the *i*th biobank (or pooled sample size across all ancestry-specific biobank summary statistics), the SSW TWAS Z-score, *Z*_*TWAS,SSW*_, is calculated as:

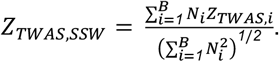

For the ancestry-unaware TWAS, we use ancestry-unaware elastic net regression models and integrate with GWAS summary statistics meta-analyzed across all ancestry groups and biobanks.

### GReX-level phenome-wide association studies (GReX-PheWAS)

Transcriptome-wide significant genes are further prioritized by performing GReX-PheWAS to categorize associations across a broad spectrum of phenotypes. Using UKBB summary statistics from European ancestry patients (Bycroft et al., 2018), we tested for GTAs for 731 phenotypes grouped into 9 categories: dermatologic, digestive, endocrine/metabolic, genitourinary, hematopoietic, musculoskeletal, neoplasms, neurological, and respiratory. Here, we illustrate GReX-PheWAS using three genes from the European-specific TWAS for asthma risk using lung tissue expression: *TAF7* (MOSTWAS model), *IL18RAP* (JTI model), and *TMEM258* (JTI model). A phenome-wide significant association was defined via Bonferroni correction 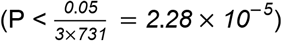.

## SUPPLEMENTAL INFORMATION

**Table S1:** *Difference in adjusted R*^*2*^ *between models trained in aligned and misaligned ancestry samples as the ancestry of the imputation sample*.

**Table S2:** *R*^*2*^ *of ancestry-specific models imputed into EUR imputation sample (training:imputation)*

**Table S3**: *R*^*2*^ *of ancestry-specific models imputed into AFR imputation sample (training:imputation)*

**Table S4**: *Difference in R*^*2*^ *between ancestry-specific and ancestry-unaware models across MAF*

**Table S5**: *Cross-validation R*^*2*^ *of ancestry-unaware models across MAF threshold*

**Table S6**: *GReX-PheWAS results for 3 representative genes from asthma meta-analytic TWAS in Europeans in GBMI that meet Bonferroni correction (P < 0*.*05/731)*

**Figure S1:** *Ratio of predictive performance of expression models in aligned versus misaligned imputation samples across AFR (left) and EUR (right) ancestry in the imputation sample*. Here, we down-sample the EUR imputation sample to match the sample size of the AFR imputation sample.

**Figure S2**: *Predictive performance of expression models in aligned and misaligned imputation samples*.

**Figure S3**: *Predictive performance of ancestry-unaware expression models compared to ancestry-specific models across 5 tissues*. Boxplot of difference in predictive performance in EUR **(A)** and AFR **(B)** samples between ancestry-aligned models and ancestry-unaware models. We consider (1) individuals of all ancestry in the training sample of the ancestry-unaware model (gold) or only EUR and AFR individuals in the training sample (grey). The red line indicates a difference of 0.

**Figure S4**: *Predictive performance of ancestry-unaware expression models across minor allele frequency thresholds*.

**Figure S5**: *TWAS Miami plots across AFR and EUR ancestry groups for asthma using whole blood gene expression models*.

**Figure S6**: *Correlation of TWAS Z-scores across ancestry-specific, individual biobank GWAS cohorts and 5 meta-analytic strategies*.

**Figure S7**: *TWAS associations across EUR and AFR ancestry groups for DFFA across 5 tissues*. The effect size is given with the point (triangle if association is transcriptome-wide significant) with a 95% confidence interval provided.

**Figure S8:** *Miami plots of DFFA local-eQTLs and GWAS signal for SNPs around DFFA*. In (A), color shows linkage disequilibrium R^2^ to lead eQTL SNP. Grey line shows a nominal P-value cutoff of 0.05 (|Z| = 1.96).

**Figure S9:** *Empirical Bayes estimates of bias and inflation in TWAS Z-scores across meta-analysis strategies*. Estimates of bias (top) and bottom (inflation) with one standard error width around the estimate are given across meta-analysis strategies. The dotted lines provide a reference for the null (0 for bias and 1 for inflation).

**Figure S10**: *Comparison of two IVW meta-analyzed Z-scores*. Vertical and horizontal dotted lines give a reference for the Bonferroni-corrected threshold for transcriptome-significance. A diagonal line is provided for reference.

**Figure S11**: *Comparison of meta-analyzed Z-scores with individual biobank TWAS Z-scores*. Ancestry-specific TWAS Z-scores for individual biobanks are shown in the top panel, colored by ancestry. Meta-analyzed Z-scores are shown in the bottom panel with shapes reflecting the different strategies. Dotted lines provide a reference for transcriptome-wide significance.

**Figure S12**: *UKBB T-PheWAS associations across 5 representative asthma-associated genes through European-only meta-analytic TWAS, grouped by phecode group*. The horizontal dotted line shows FDR-adjusted P = 0.05.

**Figure S13**: *Miami plots of UKBB T-PheWAS associations across 2 genes previously implicated through GWAS and detected in European-only meta-analytic TWAS in GBMI*.

